# Identifying the genetic constraint characteristics and biological correlates of sudden unexplained death susceptibility genes

**DOI:** 10.1101/2025.02.19.25322581

**Authors:** Junyi Lin, Chenyang Xu, Qi Shen, Ruiyang Tao, Shouyu Wang

## Abstract

Sudden unexplained death (SUD) refers to sudden death in which the cause of death could not be established after a comprehensive medico-legal investigation, including pathological/toxicological assessment and forensic investigation of the circumstances of death. In the past decade, molecular autopsy has proved to be an efficient diagnostic tool in the multidisciplinary management of SUD. Though numerous pathogenic/likely pathogenic (P/LP) variants have been identified, there is still a large proportion of SUD cases without clear molecular autopsy findings, suggesting that the genetic predisposition contributing to SUD might be more complicated than expected. In this study, we analyzed published datasets to characterize the distribution pattern of rare P/LP variants. Enrichment analyses were subsequently conducted to identify the most affected canonical pathways/biological processes and phenotypes/cell types with strongest correlation to SUD. In addition, the constraint metrics of genes harboring P/LP variants were also assessed to investigate the role of selective pressure in shaping the genetic features of SUD.

## 1. Introduction

Sudden unexplained death (SUD) constitutes a considerable portion of unexpected sudden natural death in children and young adults. Though primary arrhythmia syndromes have been proposed as the most important risk factor for SUD, its pathophysiology and genetic background need to be further elucidated. Molecular autopsy has proved to be an efficient diagnostic tool in disclosing the genetic background of SUD. By post-mortem genetic testing, pathogenic/likely pathogenic (P/LP) variants in cardiovascular and metabolic genes were shown to exist in about one-third of the SUD cases. Based on published datasets, we aim to identify the genetic constraint characteristics and biological correlates of SUD susceptibility genes.

## 2. Material studied, methods, techniques

Our analysis included 12 studies focusing on the genetics of SUD, sudden arrhythmic death syndrome (SADS), or sudden infant death syndrome (SIDS) [1-12]. Enrichment analyses were conducted using Metascape [13] to identify the most affected canonical pathways/biological processes and correlated phenotypes/cell types. In addition, a comprehensive assessment of the constraint metrics and intolerance scores of genes harboring P/LP variants was performed to investigate the role of selective pressure in shaping the genetic features of SUD. Specifically, the Loss-of-function Observed/Expected Upper bound Fraction (LOEUF) [14], Residual Variation Intolerance Score (RVIS) percentile [15], protein-coding and non-coding Genomic Evolutionary Rate Profiling (pcGERP/ncGERP) percentile [16] was respectively evaluated. Continuous variables were presented as mean ± standard error of the mean (SEM). Comparisons between two groups were made using the Mann-Whitney U test. Pairwise multiple comparisons were made using the one-way ANOVA test and Dunn’s multiple comparison post-test. All statistical tests were two-sided and a P value < 0.05 was considered statistically significant.

## 3. Results and discussion

In brief, 251 rare P/LP variants distributed in 113 different genes were identified from a total of 1272 cases. As shown in **Figure 1A**, *RYR2* was found to be the most frequently affected gene (37 P/LP variants), followed by *SCN5A* (12 P/LP variants), *TTN* (12 P/LP variants), *KCNH2* (10 P/LP variants) and *KCNQ1* (8 P/LP variants). Previous studies already concluded that dysfunction of the critical calcium/sodium/potassium channel encoding genes could lead to ion channelopathies manifested as severe inherited cardiac arrhythmias, including long QT syndrome, short QT syndrome, and Brugada syndrome [17]. In addition, the giant muscle filament titin encoding gene, *TTN*, was reported to be associated with different subtypes of cardiomyopathies [18].

**Figure 1.**
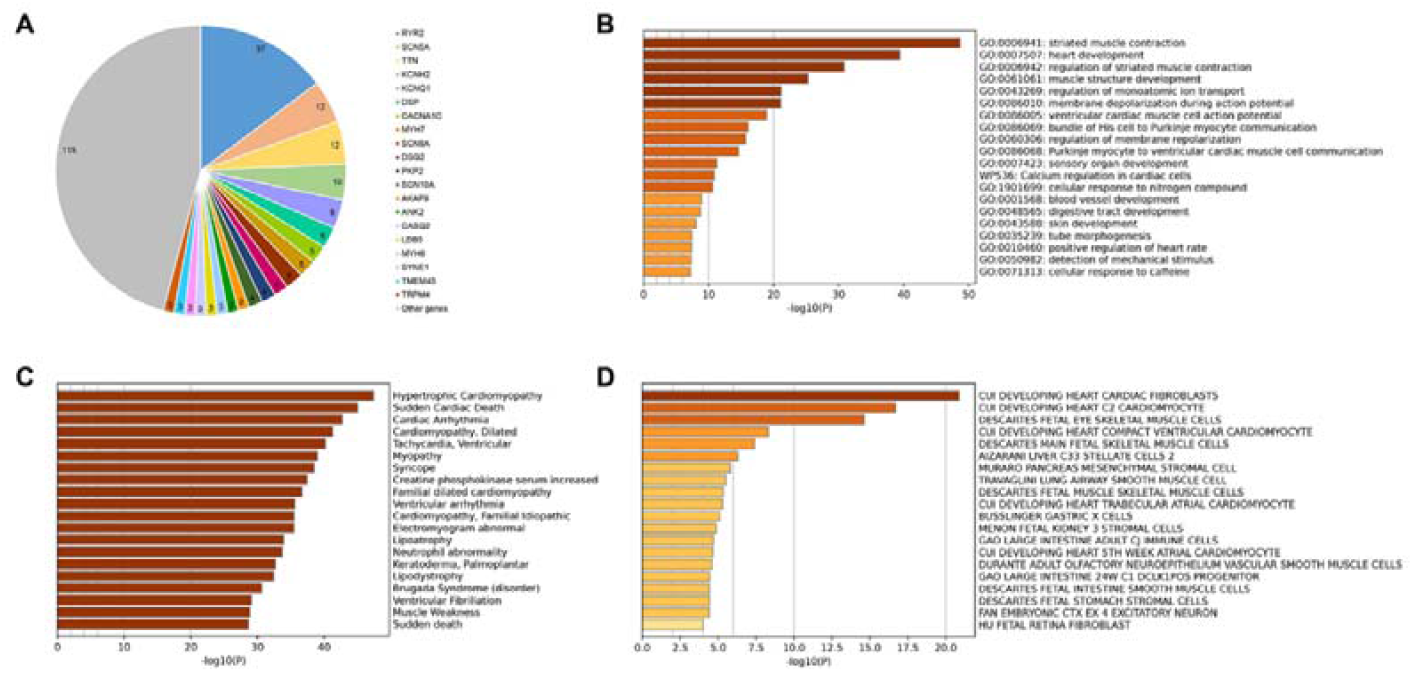
Gene-based distribution of P/LP variants and enrichment analysis results.

Among enriched pathways and processes, striated muscle contraction (GO: 0006941) was identified as the most relevant biological process (**Figure 1B**), followed by heart development (GO: 0007507) and regulation of striated muscle contraction (GO: 0006942). In terms of gene-disease association, hypertrophic cardiomyopathy was identified as the most common disease phenotype, followed by sudden cardiac death and cardiac arrhythmia (**Figure 1C**). These findings further confirmed that cardiac dysfunction is the most important pathologic basis of SUD. In addition, an enrichment analysis in Cell Type Signatures revealed that a majority of the 113 candidate genes were abundantly expressed in cardiac fibroblasts, cardiomyocytes, and fetal eye skeletal muscle cells (**Figure 1D**). These findings suggest that except for cell types known to be involved in cardiac dysfunction, disorders of other muscular organs might also be observed. Therefore, during the investigation of suspicious SUD, a thorough review of the medical history is recommended to obtain suggestive evidences supportive of molecular autopsy findings.

Regarding constraint metrics and intolerance scores, the 113 candidate genes harboring P/LP variants have consistently lower LOEUF, RVIS percentile, pcGERP percentile, and ncGERP percentile values compared with 113 random genes from GENCODE Human Release 31 (**Figure 2**). Moreover, a stepwise comparison among genes that harbor different amount of variants (Group A: 1 variant; Group B: 2-4 variants; Group C: more than 5 variants) showed genes harboring more P/LP variants have lower LOEUF, RVIS percentile, pcGERP percentile, and ncGERP percentile values (**Figure 3**). Low LOEUF scores indicate strong selection against predicted loss-of-function (pLoF) variants in a given gene, while high LOEUF scores suggest a relatively higher tolerance to inactivation. Similarly, genes with lower RVIS percentile values are more intolerant to sequence variation compared to genes with higher RVIS percentile values. In addition, lower pcGERP percentile and ncGERP percentile values indicate that the genes have relatively strong dosage sensitivities due to their highly conserved protein-coding and non-coding sequences compared to the rest of the genome. Taken together, our findings suggest that genes reported to harbor substantial P/LP variants are relatively intolerant to deleterious genetic variants in either coding or non-coding region. Due to selective pressure, variants with functional effects in these genes have higher risks of leading to severe phenotypes before reproductive age. Correspondingly, the genetic constraint characteristics could be used as an important index for the interpretation of molecular autopsy findings, especially when there is a lack of evidence regarding the genotype-phenotype correlations.

**Figure 2.**
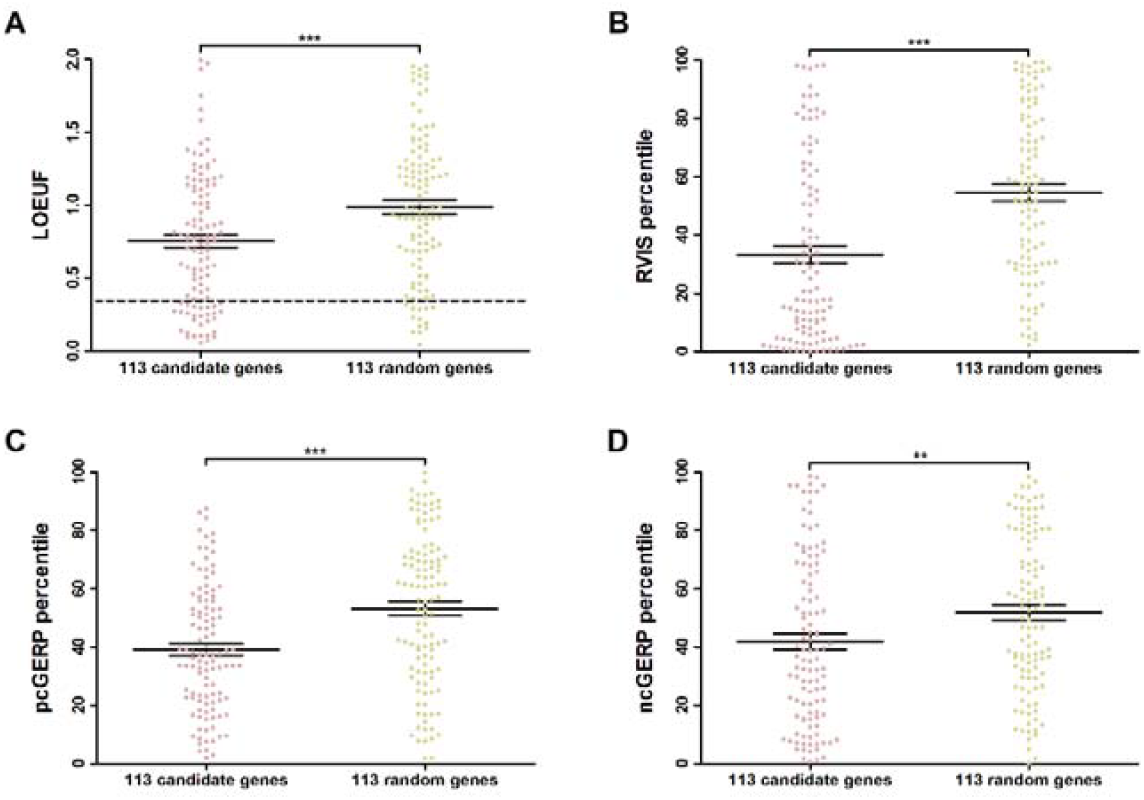
The 113 candidate genes have lower LOEUF, RVIS percentile, pcGERP and ncGERP percentile values compared with 113 random genes (^**^P ≤ 0.01; ***P ≤ 0.001).

**Figure 3.**
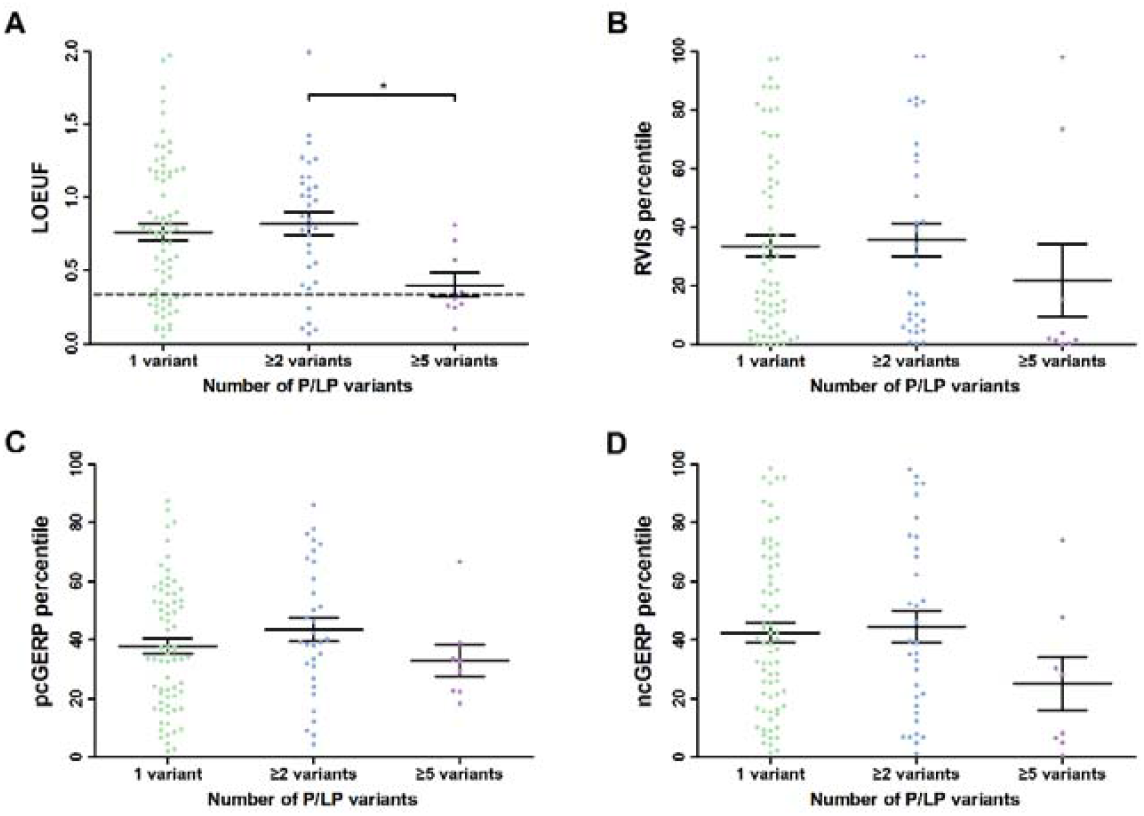
Genes harboring more P/LP variants have relatively lower LOEUF, RVIS percentile, pcGERP and ncGERP percentile values (^*^P≤0.05).

## 4. Conclusion

By analyzing published datasets from SUD/SADS/SIDS studies where a molecular autopsy was performed, the distribution pattern of 251 rare P/LP variants was characterized. Within the 113 genes harboring P/LP variants, ion channel encoding genes were found to be the most frequently affected genes. Among enriched pathways and processes, striated muscle contraction was identified as the most relevant biological process, while hypertrophic cardiomyopathy was identified as the most common disease phenotype. In addition to cardiac fibroblasts and cardiomyocytes, a majority of the candidate genes were also abundantly expressed in other cell types. More importantly, genes harboring more P/LP variants generally have lower LOEUF, RVIS percentile, pcGERP and ncGERP percentile values, suggesting that the genetic constraint characteristics might be used as an index when evaluating molecular autopsy results. In conclusion, our study provides insights into a better way of interpreting molecular autopsy findings.

## Data Availability

All data produced in the present study are available upon reasonable request to the authors

## 5. Acknowledgments

This study was supported by the National Science Foundation of China (82101971) and the Opening Project of Shanghai Key Laboratory of Crime Scene Evidence (2018XCWZK21).

